# CAIRS: A causal artificial intelligence recommendation system for digital mental health

**DOI:** 10.1101/2024.11.11.24317126

**Authors:** Mathew Varidel, Victor An, Ian B. Hickie, Sally Cripps, Roman Marchant, Jan Scott, Jacob J. Crouse, Adam Poulsen, Bridianne O’Dea, Frank Iorfino

## Abstract

Digital mental health tools have the prospect to enhance and expand access to care for those in need. Some tools provide interventional recommendations to individuals, typically using simple static rule-based systems (e.g., if-else statements) or by incorporating predictive artificial intelligence. However, interventional recommendations require a decision based on the comparison of future outcomes under different interventions, which requires causal considerations. Here we develop CAIRS, a causal artificial intelligence recommendation system that provides personalised interventional recommendations using an individual’s current presentation and the learned dynamics between domains to identify and rank intervention targets that have the greatest impact on future outcomes. Our approach was applied to longitudinal data of multiple mental health and related domains at two timepoints (1 week - 6 months from baseline) collected from a digital mental health tool. In our example, psychological distress was found to be the key influential domain that affected multiple domains (e.g., personal functioning, social connection), and thus was typically the preferred target in complex cases where multiple domains were unhealthy. Our approach is broadly applicable to recommendation contexts where causal considerations are important, and the framework could be incorporated within a live app to enhance digital mental health tools.

## Introduction

Theory suggests that mental ill-health and poor wellbeing are emergent phenomena from the interactions between symptoms within and across domains over time^1–4^. This view recognises that domains—mental health, physical health and activity, social support, personal functioning (hereafter ‘functioning’), sleep, nutrition, alcohol or other substance use—influence each other in complex ways. Furthermore, mental ill-health and wellbeing are influenced by cultural, socioeconomic, or population-level factors^5–7^. This provides a landscape of potential interventional targets that could alleviate mental ill-health and improve wellbeing. However, this landscape raises the problem of deciding between interventions, which is made difficult by; 1) the heterogeneous presentation of mental ill-health and poor wellbeing, 2) uncertainty about outcomes under different interventions, 4) timeliness, duration, and sequence of interventions, 5) comparison of multidimensional outcomes given individual-level differences in the prioritisation of different symptoms or domains, and 6) costs (monetary or otherwise) associated with interventions.

This interventional decision-making problem can be considered within a Bayesian decision-theoretic (BDT) framework^8^. In BDT the optimal interventional decision is that which maximises the expected utility of outcomes. BDT allows for the incorporation of uncertainties in components of the decision-making process (e.g., uncertainty of future outcomes under interventions). Subjective utilities can be used to account for varying individual-level multidimensional outcome preferences and perceived costs. Timing and sequencing can be addressed by conditioning current decisions on prior decisions and observations, while possible future decisions can be marginalised out.

Recommendation systems (RSs) are algorithms that filter content or actions for end-users. RSs are often built to filter content to align with an individual’s prior choices or elicited information, and have been successful for this purpose (e.g., Netflix^9^), including applications to digital mental health tools^10,11^. However, we will consider the more ambitious task of building an RS that makes interventional recommendations, by aligning recommendations with interventional decision-making considerations using a BDT framework.

This approach requires methods to predict outcomes under interventions. RSs typically use predictive artificial intelligence (AI) algorithms that estimate relationships between inputs and outcomes using conditional probabilities inferred from observational data. However, this is problematic as these relationships can be confounded in observational data, such that conditional probabilities cannot be used to predict outcomes under interventions^12^. As an alternative we will explore causal AI^13^, which includes mechanisms to estimate causal effects from observational data.

Many digital technologies aimed at improving mental health and wellbeing exist today^14–16^, but their reliance on rule-based or predictive algorithms limits their ability to provide interventional recommendations. While there has been interest in building causal AI applications for decision-support systems within healthcare generally^17,18^, there has been very little work on causal AI applications to digital mental healthcare. Furthermore, few applications of causal AI have been deployed as it is a relatively new field that requires further understanding and tools to find real-world applicability. We will show the applicability of ‘Causal AI for a RS’ (CAIRS) to rank domains as intervention targets in the mental health and wellbeing context. A schematic of CAIRS is shown in Fig. 1.

**Fig. 1.**
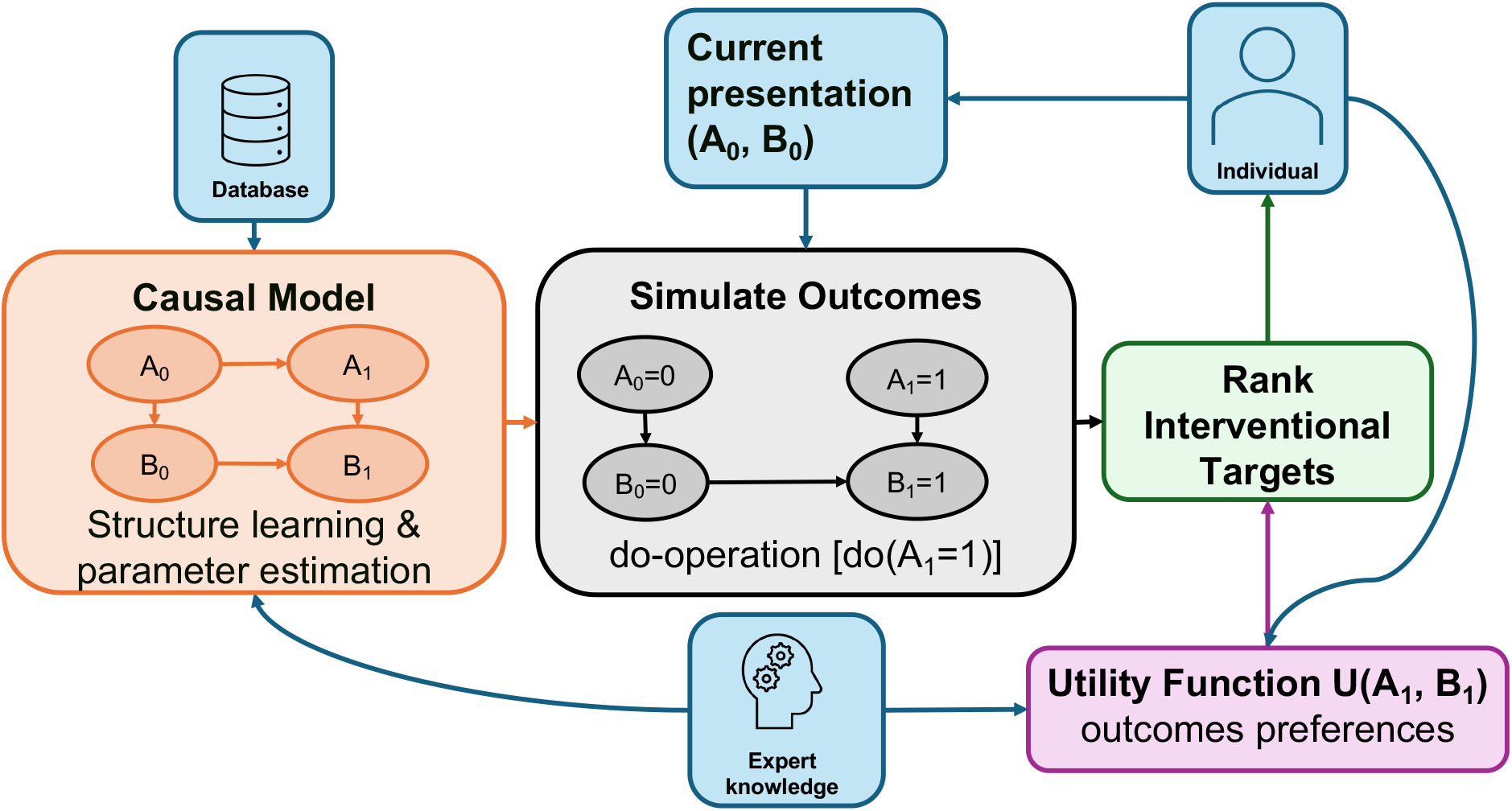
Causal artificial intelligence recommendation system (CAIRS). CAIRS uses expert knowledge and observational data to learn a causal model, which is then used to simulate outcomes under idealised interventions and displays intervention targets based on their expected utility.

## Results

### Sample characteristics

The sample comprised of 619 individuals that used the Innowell Fitness app from the original 5933 cohort. Individuals in our sample tended to have slightly higher propensity of being in the healthy category across functioning (sample, 209 [34%]; cohort, 1663 [28%]), psychological distress (sample, 326 [53%]; cohort, 2818 [48%]), nutrition (sample, 268 [43%]; cohort, 2388 [40%]), physical activity (sample, 466 [75%], cohort, 4284 [72%]), sleep (sample, 173 [28%]; cohort, 1611 [27%]), social support (sample, 180 [29%]; cohort, 1714 [29%]), and substance use (sample, 383 [62%]; cohort, 3471 [59%]). The median follow-up time was 55 days (Q1, 35 days, Q3, 96 days) with further breakdown in the appendix (Supplementary Note 1). The sample improved across a range of outcomes from baseline to follow-up where the number of individuals moving from ‘fair’ or ‘poor’ to healthy was; +45 (7.3%) for sleep, +17 (2.7%) for physical activity, +28 (4.5%) for social support, -1 (0.2%) for functioning, +50 (8.1%) for psychological distress, 0 (0.0%) for substance use, and +24 (3.9%) for nutrition. Further details in table 1.

**Table 1.** Sample characteristics. Comparison of the analysed sample (N=619; 10%) to the cohort of working individuals that have used the Innowell Fitness app (N=5933).

|  | **Healthy** | **Fair** | **Poor** |
| --- | --- | --- | --- |
| **Sleep** |  |  |  |
| Baseline (cohort), N (%) | 1611 (27%) | 3282 (55%) | 1040 (18%) |
| Baseline | 173 (28%) | 343 (55%) | 103 (17%) |
| Follow-up | 218 (35%) | 307 (50%) | 94 (15%) |
| Difference between Timepoints | 45 (7.3%) | -36 (5.8%) | -9 (1.5%) |
| **Physical activity** |  |  |  |
| Baseline (cohort) | 4284 (72%) | 294 (5.0%) | 1355 (23%) |
| Baseline | 466 (75%) | 22 (3.5%) | 131 (21%) |
| Follow-up | 483 (78%) | 24 (3.9%) | 112 (18%) |
| Difference between Timepoints | 17 (2.7%) | 2 (0.3%) | -19 (3.1%) |
| **Social support** |  |  |  |
| Baseline (cohort) | 1714 (29%) | 1675 (28%) | 2544 (43%) |
| Baseline | 180 (29%) | 169 (27%) | 267 (43%) |
| Follow-up | 208 (34%) | 205 (33%) | 203 (32%) |
| Difference between Timepoints | 28 (4.5%) | 36 (5.8%) | -64 (10%) |
| **Functioning** |  |  |  |
| Baseline (cohort) | 1663 (28%) | 1907 (32%) | 2363 (40%) |
| Baseline | 209 (34%) | 186 (30%) | 224 (36%) |
| Follow-up | 208 (34%) | 181 (29%) | 230 (37%) |
| Difference between Timepoints | -1 (0.2%) | -5 (0.8%) | 6 (1.0%) |
| **Psychological distress** |  |  |  |
| Baseline (cohort) | 2818 (48%) | 1784 (30%) | 1331 (22%) |
| Baseline | 326 (53%) | 171 (28%) | 122 (20%) |
| Follow-up | 376 (61%) | 119 (19%) | 124 (20%) |
| Difference between Timepoints | 50 (8.1%) | -52 (8.2%) | 2 (0.3%) |
| **Substance use** |  |  |  |
| Baseline (cohort) | 3471 (59%) | 1629 (28%) | 833 (14%) |
| Baseline | 383 (62%) | 158 (26%) | 78 (13%) |
| Follow-up | 383 (62%) | 158 (26%) | 78 (13%) |
| Difference between Timepoints | 0 (0.0%) | 0 (0.0%) | 0 (0.0%) |
| **Nutrition** |  |  |  |
| Baseline (cohort) | 2388 (40%) | 2106 (36%) | 1439 (24%) |
| Baseline | 268 (43%) | 213 (34%) | 138 (22%) |
| Follow-up | 292 (47%) | 206 (33%) | 121 (20%) |
| Difference between Timepoints | 24 (3.9%) | -7 (1.1%) | -17 (2.7%) |

### Structure learning

Fig. 2 is a summarisation of the posterior distribution of directed acyclic graphs (DAGs). The contemporaneous network within baseline (i.e., *A* _baseline_ → *B* _followup_) shows a dense network consistent with analysis in a partly overlapping sample^19^. There is significant uncertainty about directionality, although psychological distress was more likely to be the parent of another domain than in the opposite direction. Specifically, psychological distress being the parent of; 1) functioning was 64% compared to 36% in the opposite direction, 2) physical activity was 78% compared to 20%, 3) sleep was 80% compared to 20%, and 4) 65% to social support compared to 27%. Also, functioning was more likely to affect nutrition (79% compared to 21%).

**Fig. 2.**
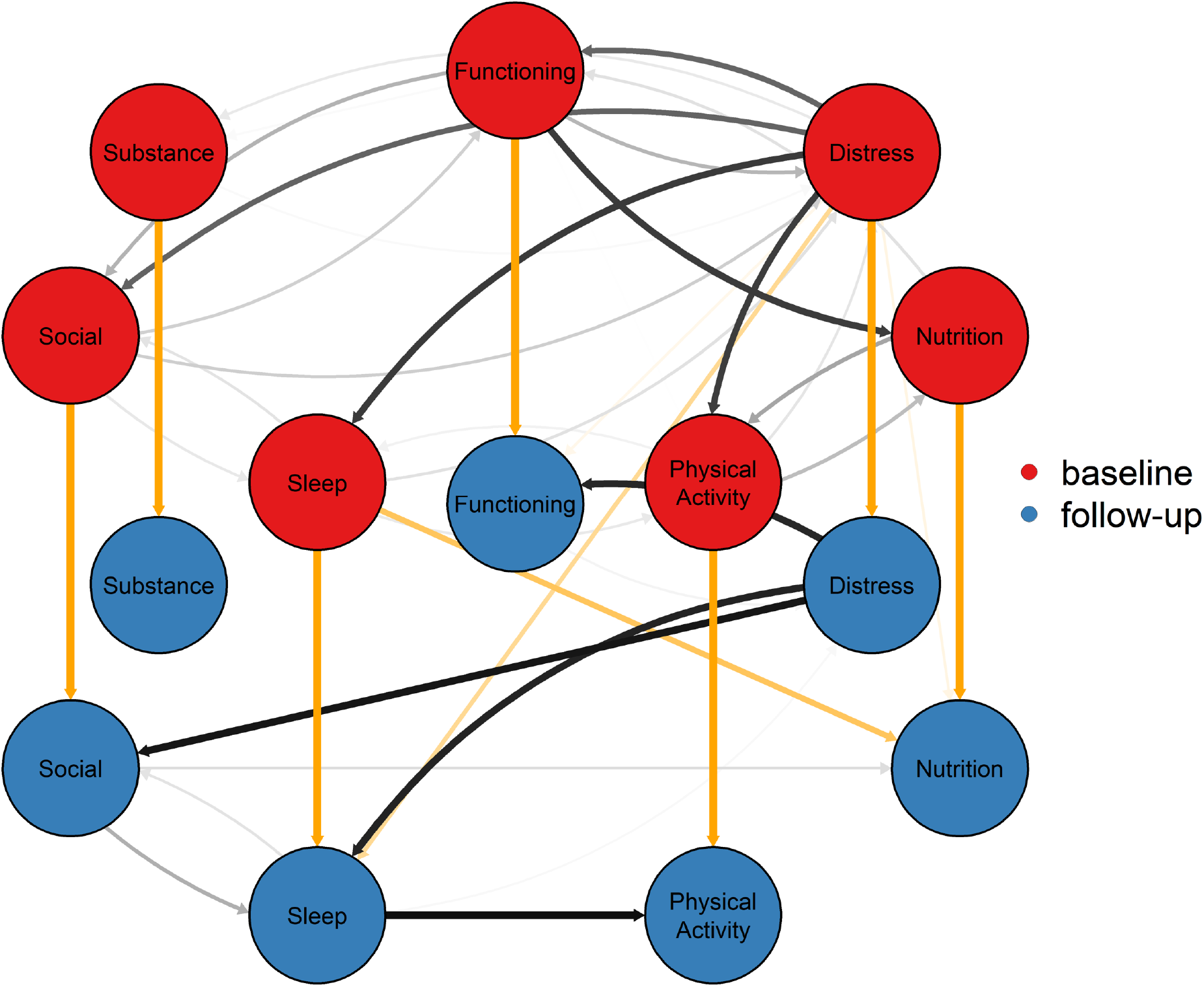
Consensus graph summarising the posterior distribution of DAGs. Edges between nodes with p_parent_>10%, with darker colours corresponding to higher probability. Yellow arrows show lagged edges whereas grey arrows show within timepoint edges.

Adding the time component helps the algorithm differentiate directionality. We found the autoregressive edges (i.e., *A*_baseline_ → *A*_followup,_) had p>99% for all domains. The lagged cross-domain edges (i.e., *A*_baseline_→ *B*_followup,_) are sleep to nutrition (p_parent_=68%) and potentially psychological distress to sleep (p_parent_=48%). Within the follow-up timepoint we found psychological distress to functioning (p_parent_=86%), social support (p_parent_=92%), and sleep (p_parent_=88%), along with sleep to physical activity (p_parent_=95%). Including indirect paths, there were paths from psychological distress to functioning (p_path_=86%), social support (p_path_=92%), sleep (p_path_=88%), and physical activity (p_path_=86%). Further details are in Supplementary Note 2.

### Treatment effects

Average treatment effects (ATEs) are shown in Fig. 3A. Intervening on psychological distress is capable of the greatest ATE, with ATE>1 (equivalent to transitioning from ‘poor’ to ‘healthy’ for one domain), when psychological distress itself was adjusted from ‘poor’ to ‘healthy’ while also affecting other domains. Interventions on other domains resulted in ATE less than one due to affects being primarily isolated to that domain.

**Fig. 3.**
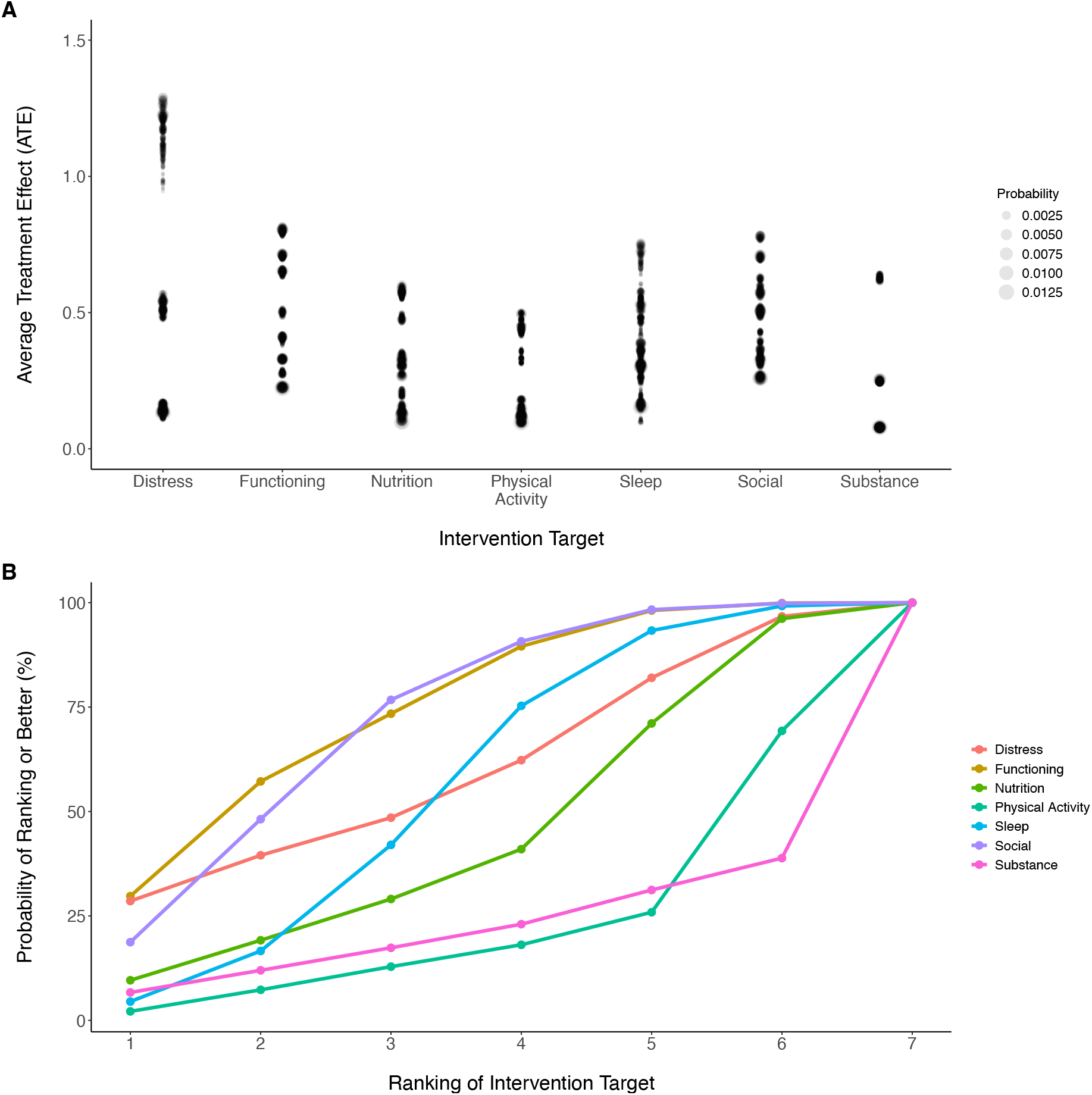
Treatment effects and recommendation targets. Panel A shows treatment effects with point size increasing with empirical probability of the baseline state. These are converted to recommendations represented in panel B, where the probability for the rank of each intervention target is marginalised over the baseline states.

### Decision analysis

We estimated a preference ranking for each domain shown in Fig. 3B. The proportion that a domain is the optimal intervention weighted in accordance with the probability of the baseline states is functioning (p_opt_=30%), psychological distress (p_opt_=29%), social support (p_opt_=18%), nutrition (p_opt_=9.6%), substance use (p_opt_=6.7%), sleep (p_opt_=4.5%), and physical activity (p_opt_=2.2%).

Typical rankings of interventional targets would affect the presentation in a system that presented the top N interventional targets. For example, the proportion that each domain would be recommended in a system that showed the top three intervention targets, would be social support (p_rec_=77%), functioning (p_rec_=73%), psychological distress (p_rec_=49%), sleep (p_rec_=42%), nutrition (p_rec_=29%), substance use (p_rec_=17%), and physical activity (p_rec_=13%).

Optimal interventional targets can be further illuminated using examples. The most common baseline presentation is everything is ‘healthy’, where the optimal interventional target was social support (EU [expected utility], 6.345, SE [standard error], 0.034) followed by functioning (EU, 6.310, SE, 0.035) which were greater than doing nothing (EU, 6.087, SE, 0.033). This is due to a regression to the mean effect, where social support (Eu [expected sub-utility], 0.764, SE, 0.010) and functioning (Eu, 0.784, SE, 0.010) tend to revert to unhealthy states with greater probability than other domains which all had Eu>0.9.

Domains that are less healthy than all other domains are typically the optimal interventional target. However, when multiple domains including psychological distress are ‘fair’ with either nutrition or physical activity as ‘poor’, psychological distress was the optimal target. For example, psychological distress is the optimal intervention target for the state (functioning=‘fair’, psychological distress=‘fair’, nutrition=‘fair’, physical activity=‘poor’, sleep=‘fair’, social=‘fair’, substance=‘healthy’), as it affects multiple domains.

Psychological distress was the optimal intervention if it was equally unhealthy to any other domain. For example, assuming the baseline state (functioning=‘poor’, psychological distress=‘poor’, nutrition=‘fair’, physical activity=‘healthy’, sleep=‘fair’, social=‘poor’, substance=‘healthy’) shown in Fig. 4A, the optimal intervention target is psychological distress (EU, 5.156, SE, 0.115), rather than functioning (EU, 4.734, SE, 0.087), or social support (EU, 4.638, SE, 0.122) despite those domains also being poor.

**Fig. 4.**
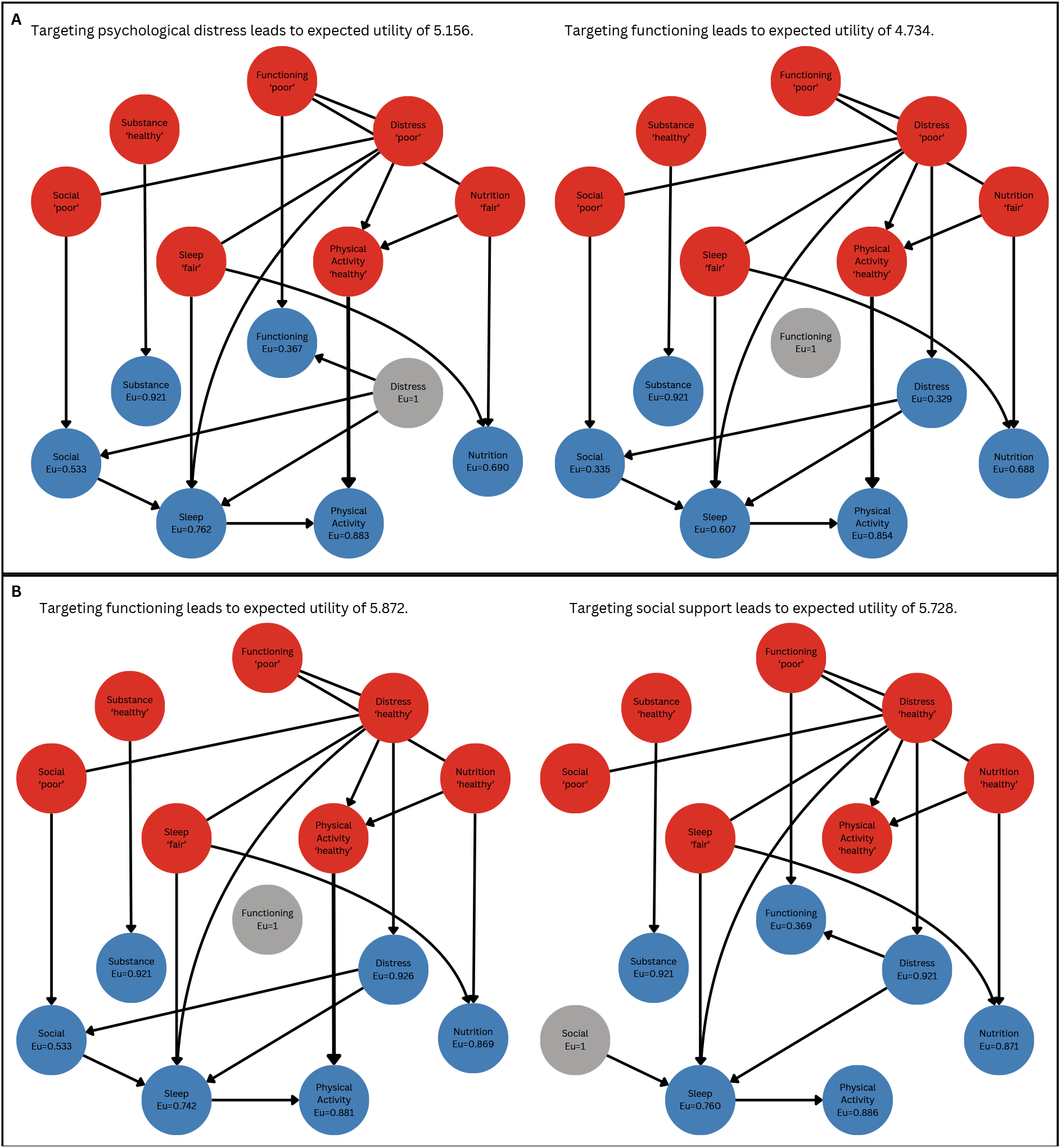
Comparison of predicted outcomes and utilities under interventions for different baseline presentations. We show edges in the *maximum a posteriori* completed partially directed acyclic graph, where undirected edges correspond to cases where DAGs with edges in either direction have the same posterior probability.

When psychological distress was ‘healthy’ and multiple domains were unhealthy, the interventional target was less certain. For example, for the state (functioning=‘poor’, psychological distress=‘healthy’, nutrition=‘healthy’, physical activity=‘healthy’, sleep=‘fair’, social=‘poor’, substance=‘healthy’) shown in Fig. 4B, the optimal interventional target was functioning (EU, 5.872, SE, 0.059) instead of social support (EU, 5.728, SE, 0.067), for similar reasons that the unhealthy functioning state persists with greater probability than social support.

We tested the sensitivity of preference rankings to utility function assumptions. Assuming a risk-neutral sub-utility function with u=(‘poor’=0, ‘fair’=0.5, ‘healthy’=1), the optimal intervention targets were psychological distress (p_opt_=28%), functioning (p_opt_=27%), social support (p_opt_=18%), sleep (p_opt_=11%), nutrition (p_opt_=7.4%), substance use (p_opt_=7.1%), and physical activity (p_opt_=1.4%). We then tested for a highly risk-averse sub-utility function u=(‘poor’=0, ‘fair’=1, ‘healthy’=1), where we found psychological distress (p=28%), functioning (p_opt_=28%), social support (p_opt_=24%), nutrition (p_opt_=8.2%), substance use (p_opt_=6.7%), physical activity (p_opt_=4.3), and sleep (p_opt_=0.9%). We also increased the weighting of psychological distress and functioning using weakly ordered domain-ranking preferences^20^, where we found distress (p_opt_=39%), functioning (p_opt_=37%), social support (p_opt_=11%), nutrition (p_opt_=5.7%), substance use (p_opt_=4.3%), sleep (p_opt_=1.9%), and physical activity (p_opt_=1.2%). Further details are in Supplementary Note 3.

## Discussion

We developed a causal artificial intelligence interventional recommendation system (CAIRS) that can be digitalised for mental healthcare technologies. Our primary contribution is to show how causal effects under interventions and expert or individual-level outcome preferences can be incorporated into an RS using BDT. Components of our approach have been explored for before. Causal AI with structure learning has been used for epidemiological purposes in mental health^21–23^. BDT has also been used for healthcare decision problems^24–26^. However, the combination of these methods with the aim of applying it to digital mental healthcare is novel.

Using our sample, we found the optimal interventional target is a function of a person’s presentation (which in this study is their state at baseline), the non-interventional transition from baseline to follow-up, the causal effects of the intervention on itself and other domains, and the utility function. To summarise our results, the optimal interventional target was; 1) the unhealthiest baseline domain with some exceptions where psychological distress is more effective than intervening on ‘poor’ nutrition or physical activity, 2) psychological distress when it is equally or more unhealthy than other domains, or 3) the domain that is more likely to transition to or persist in an unhealthy state.

These results are consistent with our prior expectations for an IRS. Intervening on the unhealthiest domain would be the conclusion of most systems. Similarly, domains that are more likely to persist in or transition to unhealthy states, suggests that intervention is required. While many IRSs may implement this latter step if known, we note that this finding was not something we considered prior to performing the analysis and is not incorporated into the current Innowell Fitness rule-based recommendations, suggesting added value from our analysis.

Our results suggest that targeting psychological distress should be preferred over other equally unhealthy domains in this population. This result is due to the causal effects that psychological distress has on functioning, social support, sleep, and physical activity mediated by sleep. This is consistent with current understanding that mental ill-health affects multiple domains, including work with respect to absenteeism and productivity^27,28^, sleeping patterns for example due to rumination^29,30^, and social connection due to social anxiety^31^. Thus, direct interventions on psychological distress are expected to have wide-ranging effects.

Comparing multidimensional outcomes in accordance with expert or individual preferences is a challenging task that BDT provides a framework to computationally encode within an IRS. Some possibilities of varying outcome preferences were explored by adjusting the sub-utility function to account for different risk-aversity and changing the domain weightings, which corresponded to slight recommendation changes. Further adjustments in the utility function are possible and this utility framework will become increasingly important as we increase the number of domains or investigate symptoms, as it’s unlikely that all relevant domains or symptoms would be considered equally important by all individuals or experts.

Incorporating varying personalised outcome preferences in a deployed app would require a utility elicitation mechanism. There are many utility elicitation methods available^20^, some of which are not overly burdensome, such as eliciting weakly ordered preference for domains, while BDT provides mechanisms to stop utility elicitation when there is enough information to make a recommendation^32^. With that said, our analysis doesn’t address the feasibility of eliciting individualised utilities in real-world applications, which should be considered for further research.

BDT could find more applicability elsewhere within digital health applications. For example, we may want to balance mental health and wellbeing outcomes with other considerations, such as user engagement which is often poor in mental health apps^33–35^. It could also be applied to other causal inference or predictive frameworks, such as undirected networks where intervention targets have been explored^36–39^, but often assume that all domains or symptoms are equally important.

Our approach is in its early stages. The current system only generates a recommendation based on an individual’s baseline presentation. Future work will account for ongoing observations by incorporating feedback mechanisms to adapt the structural causal model (SCM) to individuals over time. Furthermore, moving from interventional targets to interventions is vital, and raises new complications as interventions often act on multiple variables simultaneously^40^, may act on mediating variables that control the relationship between variables^41^, and have associated costs (monetary or otherwise). Plus, maintaining a healthy state is different to improving the state, and thus different interventions will be required.

Our results rely on causal interpretations of the inferred DAGs. Assumptions must hold for this to be true, including that we have all relevant confounders and colliders^42^. For the causal effect estimations to be valid, this surmounts to the assumption that other factors such as sociodemographics, historical values prior to baseline, or other domains have a negligible causal effect on follow-up variables beyond the effect that they have on the observed variables. These assumptions may not hold, and should be tested more thoroughly, including improving the recording of potential confounders within the Innowell Fitness app.

The true causal paths are probably more complex than suggested in this work as more causal effects have been found in other contexts between the domains that we have studied. The data is likely underpowered to determine all causal paths. Also, within timepoint cyclic causal paths will be missing, which will require the incorporation of recent methodological developments^43^.

In summary, interventional recommendation algorithms require careful incorporation of causal consideration and decision-making principles. Important aspects are the estimation of outcomes under interventions requiring causal modelling, assignment of appropriate utilities to align recommendations with outcome preferences, and consideration of uncertainty. These considerations can be incorporated into computational models using causal AI and Bayesian decision theory.

## Methods

### Ethics

This research activity has been deemed exempt from ethics review under Section 5.1.17(a) of the Australian National Health and Medical Research Council National Statement on Ethical Conduct in Human Research (2023) by The University of Sydney Human Research Ethics Committee.

### Study design

This retrospective study was conducted using data collected from the Innowell Fitness app (N=5933). Individuals who reported negative minutes spent doing any physical activity were excluded (n=55). Otherwise, we included all individuals that had at least one follow-up from 1 week to 6 months after baseline (n=619).

### Procedures

The Innowell Fitness app is a digital technology used by adults for the assessment, management, and monitoring of their mental health and wellbeing^19^. It is available on mobile and computer devices, and includes; (1) self-report assessments about each domain of a person’s mental fitness; (2) actionable insights about each domain of mental fitness; (3) personalised (rule-based) recommendations with evidence-based strategies and resources to understand and manage mental fitness; and (4) a goal setting and tracking tool which provides people with habit-forming activities designed to improve their mental fitness. The development of this tool involved a team of psychologists, psychiatrists, mental health research experts, and those with lived experience, who selected items that measure various components of mental fitness and collated relevant evidence-based strategies and resources.

The measures and domains assessed include; (1) social support, using three items from the Schuster’s Social Support Scale^44^; (2) personal functioning (referred throughout as ‘functioning’), using three items about educational and employment engagement and achievement^45^; (3) psychological distress, using the Kessler-6^46^ scale for psychological distress; (4) sleep, using four sleep items^47–49^, including feeling refreshed after sleep, trouble falling asleep and subjective energy; (5) physical activity, four items from the International Physical Activity Questionnaire^50^ measuring time spent walking, doing moderate exercise, doing vigorous exercise, and being sedentary; (6) alcohol and other substance use, using three items about tobacco, alcohol, and other substance use^51^; and (7) nutrition, using two items about typical composition and portion size of their diet^52^. These individual items detailed in Supplementary Note 4, are combined to construct domains of interest that are categorised as either ‘poor’, ‘fair’, or ‘healthy’.

### Statistical analysis

Statistical modelling and analyses were performed in R version 4.3.3. Causal inference was performed in the structural causal modelling (SCM) framework^12^. An SCM is described by a set of variables, a set of functions relating the variables, and a causal structure represented by a DAG that indicates the directionality of causal influence using arrows between random variables. We performed Bayesian inference to infer the posterior distribution of DAGs, where we aimed to sample from the posterior distribution assuming a uniform prior over DAGs, only excluding DAGs with arrows that go backwards in time. Posterior sampling was achieved using an implementation of the Partition Markov chain Monte Carlo (PMCMC) scheme^53,54^. PMCMC samples from the space of partitions, where a partition is a set of weakly ordered nodes that represents multiple DAGs (e.g., DAG *A* ← *B* → *C* → *D* is represented by the partition {{*B*}, {*A, C*}, {*D*}}). To return to the partition space, a DAG is sampled given a partition in accordance with its posterior probability. The sampling procedure was run across eight chains and checked for convergence and resolution (Supplementary Note 5). We used the Bayesian Gaussian equivalent score function to retain the ordinal information of the random variables.

Simulating outcomes given a DAG is performed by constructing a Bayesian network (BN)^12,55^. A BN assumes nodes take categorical values and relates a variable to its parent variables using conditional probability tables. A BN was constructed per posterior sample by passing the DAG and observed data to the gRain library^56^, which estimates the conditional probability tables using a *maximum a posteriori* estimate. Simulating an outcome given an observed baseline state is performed by setting each baseline node with the values of the observed baseline state and then simulating the follow-up state. This corresponds to doing ‘nothing’ below, which is shorthand for simulating a follow-up state given no intervention. The interventional do-operation is used to simulate outcomes given idealised interventions^12,55^. This is performed by ‘mutilating’ the BN by removing all edges into an interventional node, setting the interventional node state to ‘healthy’, and then setting the states of the baseline nodes equal to the given baseline state. Note that this intervention acts on a domain at follow-up.

We assign numerical values to outcomes, which are referred to as utilities in decision theory, to make comparisons between idealised interventions. For our primary assumption, we assign the sub-utility value for outcomes as u=(‘poor’=0, ‘fair’=0.75, healthy=1), which corresponds to a moderately risk-averse sub-utility function. The utility function is then an equal-weighted sum of the sub-utility values across domains, thus assuming no preference between domains, and ensuring that our focus is on overall wellbeing. The utility ranges from zero when all domains are ‘poor’ to seven when all domains are ‘healthy’. We use the expected utility principle to order intervention targets^8^, where we assume that an intervention target *A* is preferred to *B* when the expected utility (EU) for performing an idealised intervention on *A* is greater than on *B*. In our sensitivity analysis, we investigated a risk-neutral sub-utility function with u=(‘poor’=0, ‘fair’=0.5, healthy=1), a risk-averse sub-utility function u=(‘poor’=0, ‘fair’=1, healthy=1), and up-weighting psychological distress and functioning compared to all other domains. Further detail about the BDT framework is provided in Supplementary Note 6.

We report the average treatment effect (ATE) conditional on the baseline state. This is calculated as the difference in the expected utility between doing an idealised intervention compared to doing nothing.

## Data availability

Data used for this study is available from the corresponding author on reasonable request.

## Code availability

All relevant code this paper can be found at https://github.com/VictorytA/causalintervention.

## Acknowledgements

This work was supported by the Medical Research Future Fund National Critical Research Infrastructure Grant (MRFCRI000279), and NHMRC Australia Fellowship (No. 511921 awarded to I.B.H.). M.V. was supported by philanthropic funding from The Johnston Fellowship and from other donor(s) who are families affected by mental illness who wish to remain anonymous. I.B.H. is supported by an NHMRC L3 Investigator Grant (GNT2016346). J.J.C. was supported by a NHMRC Emerging Leadership Fellowship (GNT2008196). F.I. was supported by an NHMRC EL1 Investigator Grant (GNT2018157).

## Author contributions

M.V. and F.I. conceptualised the study. M.V., V.A., S.C., and R.M. developed the software and methodologies. M.V., V.A., and F.I. wrote the first draft. Draft reviewing and editing were performed by I.B.H., S.C., R.M., J.S., J.J.C., B.O., and A.P.. All authors had full access to the data in the study and had final responsibility for the decision to submit for publication. M.V. and V.A. have directly accessed and verified the data.

## Competing interests

I.B.H. is the Co-Director, Health and Policy at the Brain and Mind Centre (BMC) University of Sydney, Australia. The BMC operates an early-intervention youth service at Camperdown under contract to headspace. I.B.H. has previously led community-based and pharmaceutical industry-supported (Wyeth, Eli Lily, Servier, Pfizer, AstraZeneca, Janssen Cilag) projects focused on the identification and better management of anxiety and depression. I.B.H. is the Chief Scientific Adviser to, and a 3.2% equity shareholder in, InnoWell Pty Ltd which aims to transform mental health services through the use of innovative technologies. All other authors declare no conflict of interest. All other authors declare no financial or non-financial competing interests.

## Notes

### Summary of Updates

Corrected order numbering of supplementary tables and figures.

