## Supplementary Material for "CAIRS: A causal artificial intelligence recommendation system for digital mental health"

**Supplementary Information**

### Supplementary Note 1. Distribution of follow-up times

**Supplementary Table 1.** Distribution of follow-up times across the population.

| **Days from baseline to follow-up** | N (%) |
|---|---|
| 7 - 30 | 125 (20%) |
| 30 - 60 | 214 (35%) |
| 60 - 90 | 104 (17%) |
| 90 - 120 | 74 (12%) |
| 120 - 150 | 49 (7.9%) |
| 150 - 180 | 53 (8.6%) |

### Supplementary Note 2. Structure learning statistics

We provide the numerical detail for the structure learning results within Supplementary Tables 2-6.

| | | Base | | | | | | |
|---|---|---|---|---|---|---|---|---|
| | | Functioning | Distress | Nutrition | Physical | Sleep | Social | Substance |
| Base | Functioning | 0 | 0.36 | 0.79 | 0.10 | 0.02 | 0.37 | 0.12 |
| | Distress | 0.64 | 0 | 0.04 | 0.78 | 0.80 | 0.65 | 0.18 |
| | Nutrition | 0.21 | 0.02 | 0 | 0.42 | 0.02 | 0.02 | 0.02 |
| | Physical | 0.07 | 0.20 | 0.33 | 0 | 0.14 | 0 | 0.03 |
| | Sleep | 0.02 | 0.20 | 0.02 | 0.20 | 0 | 0.20 | 0.04 |
| | Social | 0.27 | 0.27 | 0.02 | 0.01 | 0.18 | 0 | 0.02 |
| | Substance | 0.06 | 0.12 | 0.02 | 0.04 | 0.03 | 0.02 | 0 |

**Supplementary Table 2|** The posterior probability that a domain is a parent of another domain at baseline. Where the direction of dependency is defined as row → column.

| | | Follow-up | | | | | | |
|---|---|---|---|---|---|---|---|---|
| | | Functioning | Distress | Nutrition | Physical | Sleep | Social | Substance |
| Base | Functioning | 1 | 0.09 | 0.02 | 0.02 | 0.02 | 0.02 | 0.02 |
| | Distress | 0.18 | 1 | 0.20 | 0.02 | 0.48 | 0.10 | 0.02 |
| | Nutrition | 0.02 | 0.02 | 1 | 0.02 | 0.03 | 0.02 | 0.02 |
| | Physical | 0.02 | 0.02 | 0.05 | 1 | 0.06 | 0.02 | 0.02 |
| | Sleep | 0.04 | 0.06 | 0.68 | 0.03 | 1 | 0.02 | 0.05 |
| | Social | 0.02 | 0.07 | 0.02 | 0.02 | 0.02 | 1 | 0.02 |
| | Substance | 0.02 | 0.03 | 0.03 | 0.02 | 0.02 | 0.02 | 1 |

**Supplementary Table 3|** The posterior probability that a domain at baseline is a parent of another domain at follow-up. Where the direction of dependency is defined as row → column.

| | | Follow-up | | | | | | |
|---|---|---|---|---|---|---|---|---|
| | | Functioning | Distress | Nutrition | Physical | Sleep | Social | Substance |
| Follow-up | Functioning | 0 | 0.14 | 0.04 | 0.01 | 0.01 | 0.03 | 0.04 |
| | Distress | 0.86 | 0 | 0.10 | 0.03 | 0.88 | 0.92 | 0.02 |
| | Nutrition | 0.03 | 0.02 | 0 | 0.03 | 0.01 | 0.03 | 0.03 |
| | Physical | 0.01 | 0.03 | 0.07 | 0 | 0.02 | 0.03 | 0.03 |
| | Sleep | 0.03 | 0.12 | 0.05 | 0.95 | 0 | 0.20 | 0.03 |
| | Social | 0.03 | 0.08 | 0.22 | 0.08 | 0.38 | 0 | 0.03 |
| | Substance | 0.03 | 0.03 | 0.03 | 0.04 | 0.01 | 0.03 | 0 |

**Supplementary Table 4|** The posterior probability that a domain is a parent of another domain at follow-up. Where the direction of dependency is defined as row → column.

| | | Follow-up | | | | | | |
|---|---|---|---|---|---|---|---|---|
| | | Functioning | Distress | Nutrition | Physical | Sleep | Social | Substance |
| Base | Functioning | 1 | 0.57 | 0.93 | 0.78 | 0.67 | 0.71 | 0.37 |
| | Distress | 1 | 1 | 0.98 | 0.99 | 0.99 | 1 | 0.48 |
| | Nutrition | 0.33 | 0.25 | 1 | 0.57 | 0.34 | 0.33 | 0.19 |
| | Physical | 0.36 | 0.29 | 0.57 | 1 | 0.42 | 0.38 | 0.21 |
| | Sleep | 0.46 | 0.38 | 0.84 | 0.97 | 1 | 0.58 | 0.27 |
| | Social | 0.56 | 0.44 | 0.64 | 0.73 | 0.69 | 1 | 0.26 |
| | Substance | 0.29 | 0.24 | 0.32 | 0.36 | 0.30 | 0.30 | 1 |

**Supplementary Table 5|** The posterior probability that there is a path from a domain at baseline to a domain at follow-up. Where the path direction is defined as row → column.

| | | Follow-up | | | | | | |
|---|---|---|---|---|---|---|---|---|
| | | Functioning | Distress | Nutrition | Physical | Sleep | Social | Substance |
| Follow-up | Functioning | 0 | 0.14 | 0.11 | 0.17 | 0.16 | 0.17 | 0.05 |
| | Distress | 0.86 | 0 | 0.40 | 0.86 | 0.88 | 0.92 | 0.13 |
| | Nutrition | 0.05 | 0.03 | 0 | 0.07 | 0.04 | 0.05 | 0.03 |
| | Physical | 0.05 | 0.04 | 0.09 | 0 | 0.04 | 0.07 | 0.04 |
| | Sleep | 0.16 | 0.12 | 0.19 | 0.95 | 0 | 0.28 | 0.08 |
| | Social | 0.12 | 0.08 | 0.27 | 0.44 | 0.41 | 0 | 0.06 |
| | Substance | 0.06 | 0.04 | 0.06 | 0.09 | 0.05 | 0.06 | 0 |

**Supplementary Table 6|** The posterior probability that there is a path from a domain at follow-up to another domain at follow-up. Where the path direction is defined as row → column.

### Supplementary Note 3. Utility function sensitivity analysis

We present sensitivity to our utility functions here. We tested a risk-neutral sub-utility function $u = ('\text{poor}' = 0, '\text{fair}' = 0, '\text{healthy}' = 1)$ shown in Supplementary Figure 1 and risk-averse function $u = ('\text{poor}' = 0, '\text{fair}' = 0, '\text{healthy}' = 1)$ shown in Supplementary Figure 2.

For the domain weighted utility function, we assumed a weakly-order preference in domains where psychological distress and functioning were ranked greater than all other domains. The ranks are defined in a vector $h = (h_1, h_2, \dots, h_7)$ with the the unnormalized weight for the $j$-th domain given by $w_j = (\sqrt{2})^{h_j}$ which is normalised by dividing by $W = \sum_{j=1}^{7} (\sqrt{2})^{h_j}$ [5,6]. We assigned $h_j = 2$ for psychological distress and functioning, whereas all other domains were assigned $h_j = 1$. The total utility is defined as $U = 7 \times \Sigma_{j=1}^{7} w_j u(Z_i^j)/W$, where the multiplication by 7 maps the utility range to $U \in [0,7]$, which is consistent with the other utility function assumptions. The results for this utility function assumptions are shown in Supplementary Figure 3.

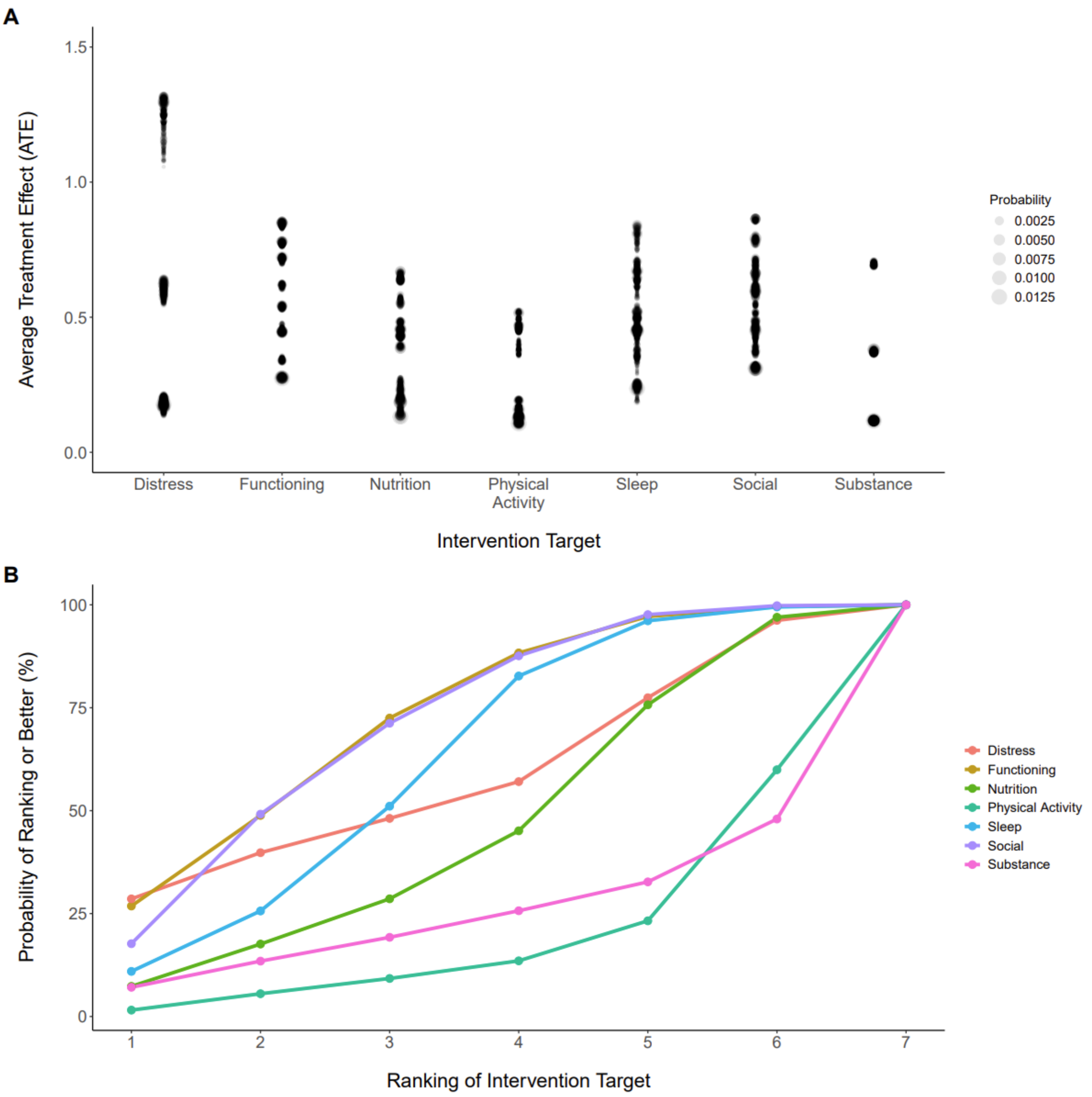

**Supplementary Fig. 1|** Treatment effects (A) and preference rankings (B) for a risk-neutral sub-utility function with 'fair' set as 0.5.

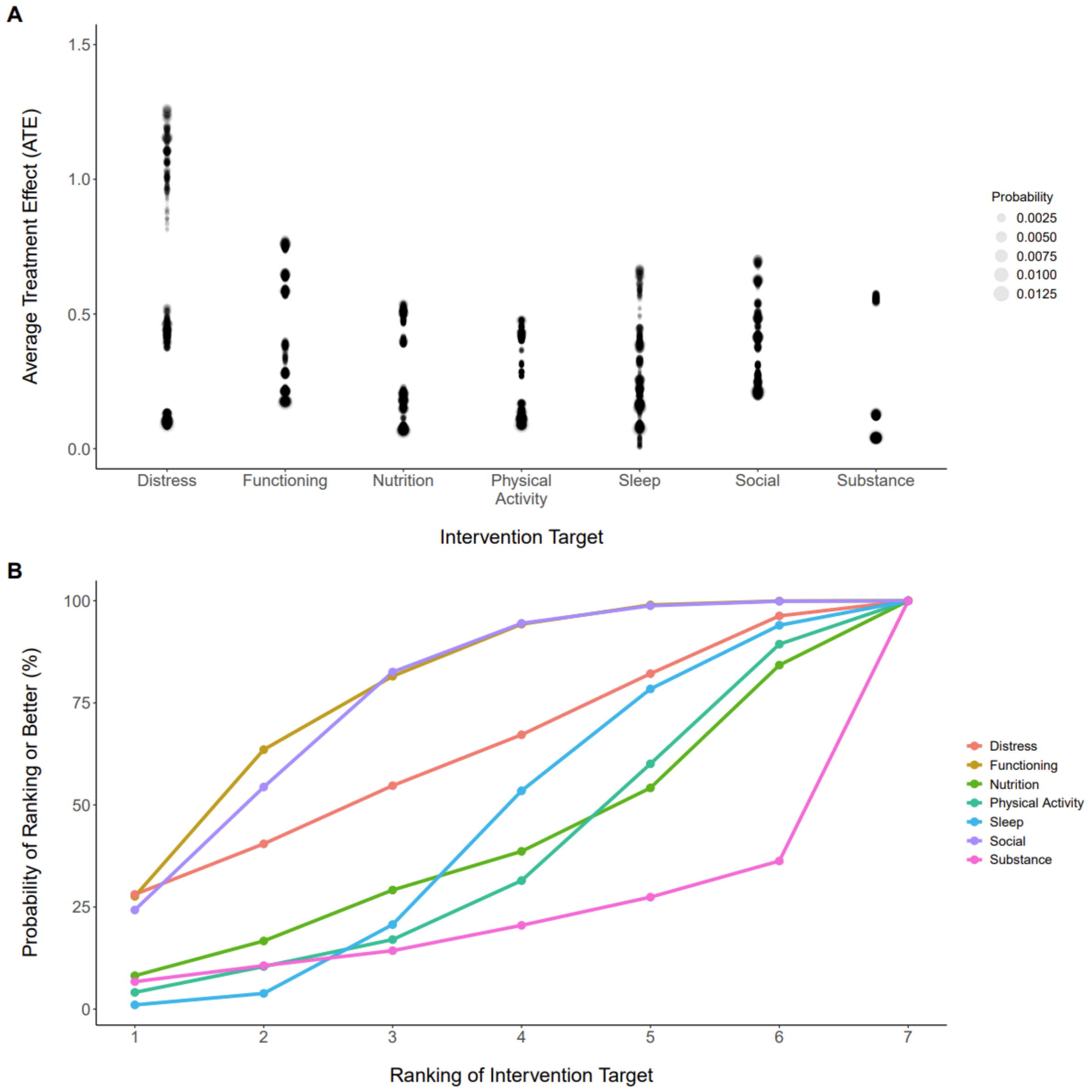

**Supplementary Fig. 2|** Treatment effects (A) and preference rankings (B) for a risk-averse sub-utility function with 'fair' set as 1.

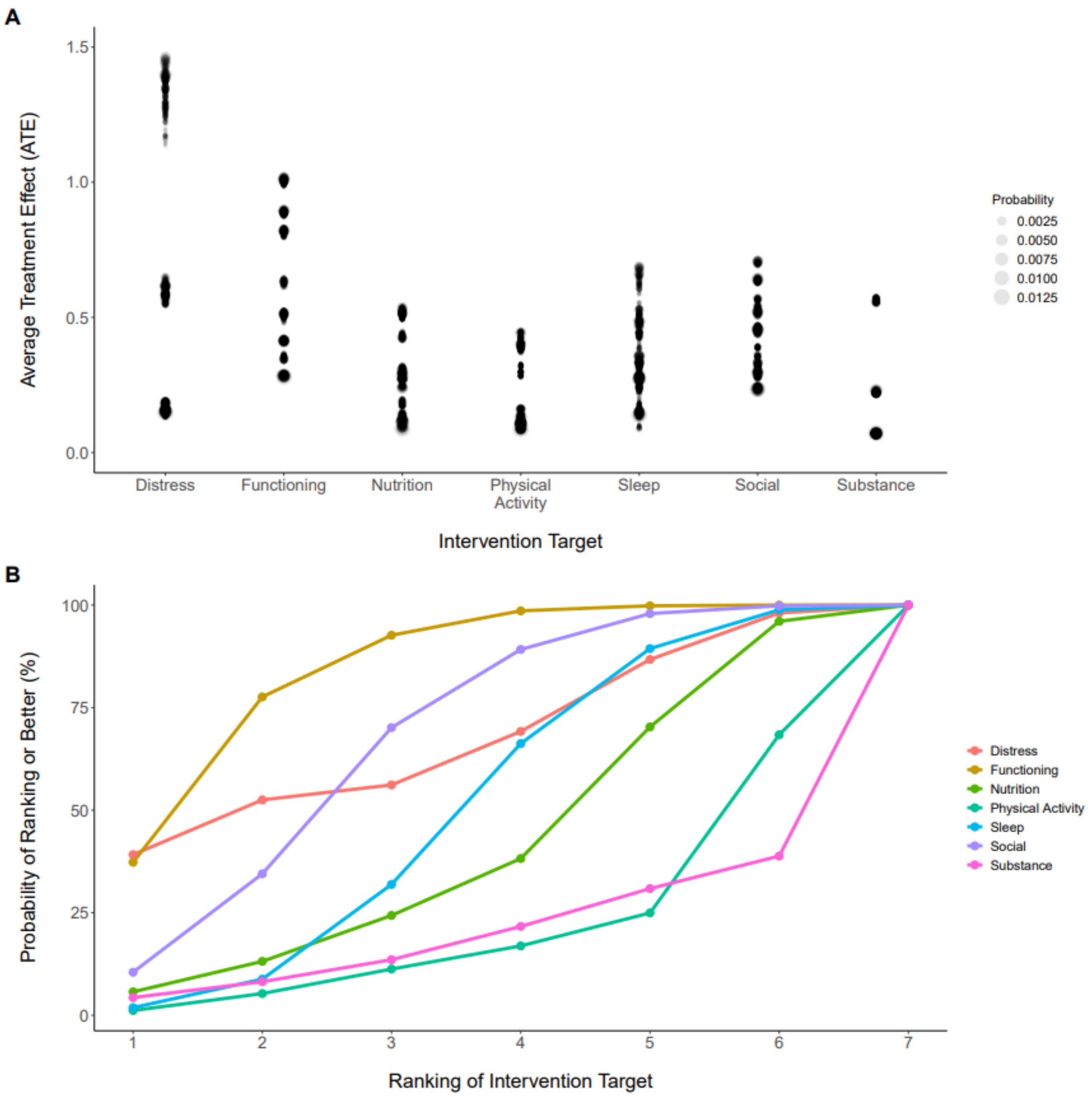

**Supplementary Fig. 3|** Treatment effects (A) and preference rankings (B) when the psychological distress and functioning domains are weighted higher than other domains.

### Supplementary Note 4. Innowell Fitness questionnaire

| Domain | Question | Answer Options |
|---|---|---|
| Sleep and circadian rhythms | How many hours of sleep did you usually get a night? | <= 6 hours |
| | | 6.5-7.5 hours |
| | | 8-10 hours |
| | | 10.5-11.5 hours |
| | | >= 12 hours |
| | How often have you felt really refreshed when waking in the morning? | Never |
| | | 1 day/week |
| | | 2-3 days/week |
| | | 4-5 days/week |
| | | 6-7 days/week |
| | Did you often have trouble falling asleep (more than 30 minutes) or wake up frequently throughout the night (>= 3 times)? | Never |
| | | 1 day/week |
| | | 2-3 days/week |
| | | 4-5 days/week |
| | | 6-7 days/week |
| | How often did you feel that you had little or no energy during the day? | Never |
| | | 1 day/week |
| | | 2-3 days/week |
| | | 4-5 days/week |
| | | 6-7 days/week |
| Physical activity | Over the past week how much time did you spend walking? | User enters hours and minutes. |
| | Over the past week how much time did you spend doing moderate physical activities? | User enters hours and minutes. |
| | Over the past week how much time did you spend doing vigorous physical activities? | User enters hours and minutes. |
| | Over the past week how much time did you spend sitting or lying down on an average weekday? | User enters hours and minutes. |
| Social connection | When you think about the amount of contact you had with your friends and/or family, do you think it was: | Too much contact |
| | | About the right amount of contact |
| | | Not enough contact |
| | How often did your friends and/or family make you feel cared for? | Never |
| | | Rarely |
| | | Sometimes |
| | | Often |
| | I felt a sense of mutual connection with my family, friends, community and culture. | Strongly agree |
| | | Agree |
| | | Neither agree nor disagree |
| | | Disagree |

| | | |
|---|---|---|
| | | Strongly disagree |
| Functioning | How many days in total were you able to carry out your usual daily activities fully? | 0 |
| | | 1 |
| | | 2 |
| | | 3 |
| | | 4 |
| | | 5 |
| | | 6 |
| | | 7 |
| | How many hours were you engaged in education, training, employment, or a designated carer role? | Wasn’t in education, training, employment, or a designated carer role |
| | | Less than 5 hours |
| | | 5 to 14 hours |
| | | 15 to 29 hours |
| | | 30 hours or more |
| | How often did you achieve the things you wanted to? | Never |
| | | Rarely |
| | | Sometimes |
| | | Often |
| Psychological distress | Over the past week, roughly how often did you feel nervous? | All of the time |
| | | Most of the time |
| | | Some of the time |
| | | A little of the time |
| | | None of the time |
| | Over the past week, roughly how often did you feel hopeless? | All of the time |
| | | Most of the time |
| | | Some of the time |
| | | A little of the time |
| | | None of the time |
| | Over the past week, roughly how often did you feel restless or fidgety? | All of the time |
| | | Most of the time |
| | | Some of the time |
| | | A little of the time |
| | | None of the time |
| | Over the past week, roughly how often did you feel so depressed that nothing could cheer you up? | All of the time |
| | | Most of the time |
| | | Some of the time |
| | | A little of the time |
| | | None of the time |
| | Over the past week, roughly how often did you feel that everything was an effort? | All of the time |
| | | Most of the time |
| | | Some of the time |
| | | A little of the time |

| | | |
|---|---|---|
| | | None of the time |
| | Over the past week, roughly how often did you feel worthless? | All of the time |
| | | Most of the time |
| | | Some of the time |
| | | A little of the time |
| | | None of the time |
| Substance Use | How often have you used tobacco products? | Never |
| | | Once or twice |
| | | Daily or almost daily |
| | How often did you consume more than four standard drinks of alcohol on one occasion? | Never |
| | | Once or twice |
| | | Daily or almost daily |
| | How often have you used other substances? | Never |
| | | Once or twice |
| | | Daily or almost daily |
| | Have you considered (or has a friend, relative or someone else suggested) improving your health and wellbeing by reducing alcohol or substance consumption? | No, I don’t want or need to cut down |
| | | Possibly, I’m thinking about it |
| | | Yes, I’m currently trying to cut down |
| | | A friend or relative or someone else has suggested it |
| Nutrition | Which of the following best describes your typical main meal? | Mostly vegetables, with some protein and carbohydrates |
| | | An even mix of vegetables, protein, and carbohydrates |
| | | Mostly carbohydrates, with some meat and vegetables |
| | | Mostly meat, with some vegetables and carbohydrates |
| | | A mix of meat and carbs, with little to no vegetables |
| | Which of the following pictures is closest to the serving size you usually consumed for a main meal? | Choice of serving sizes |

**Supplementary Table 7|** Questions for each domain.

### Supplementary Note 5. Summary of the posterior sampling

The sampling algorithm was run across eight chains where each chain started from a unique location. The starting location per chain was arrived at by sampling a DAG uniformly from the unconstrained space of DAGs with nodes consistent with our data, and then edges were removed that were inconsistent with the blacklist. Each chain was run for 400,000 iterations with the initial 1000 iterations for each chain removed as burn-in. Additionally, we thinned the chains to retain every 10$^{th}$ iteration resulting in a total of 39,900 samples for each chain.

We checked the convergence of the chains for the log posterior for the partitions and DAG. The split-$\hat{R}$ convergence statistic[1,2] for the log posterior of the partitions was consistent with the chains converging ($\hat{R}_P$, 1.003) with an effective sample size of $S_{\text{eff}} = 7{,}943$. For the log posterior of the DAGs, the split-$\hat{R}$ convergence statistic was also consistent with the chains converging ($\hat{R}_D$, 1.001) and the effective sample size was $S_{\text{eff}} = 18{,}396$. Qualitative checks of the trace plots were also consistent with convergence (see Supplementary Figure 1).

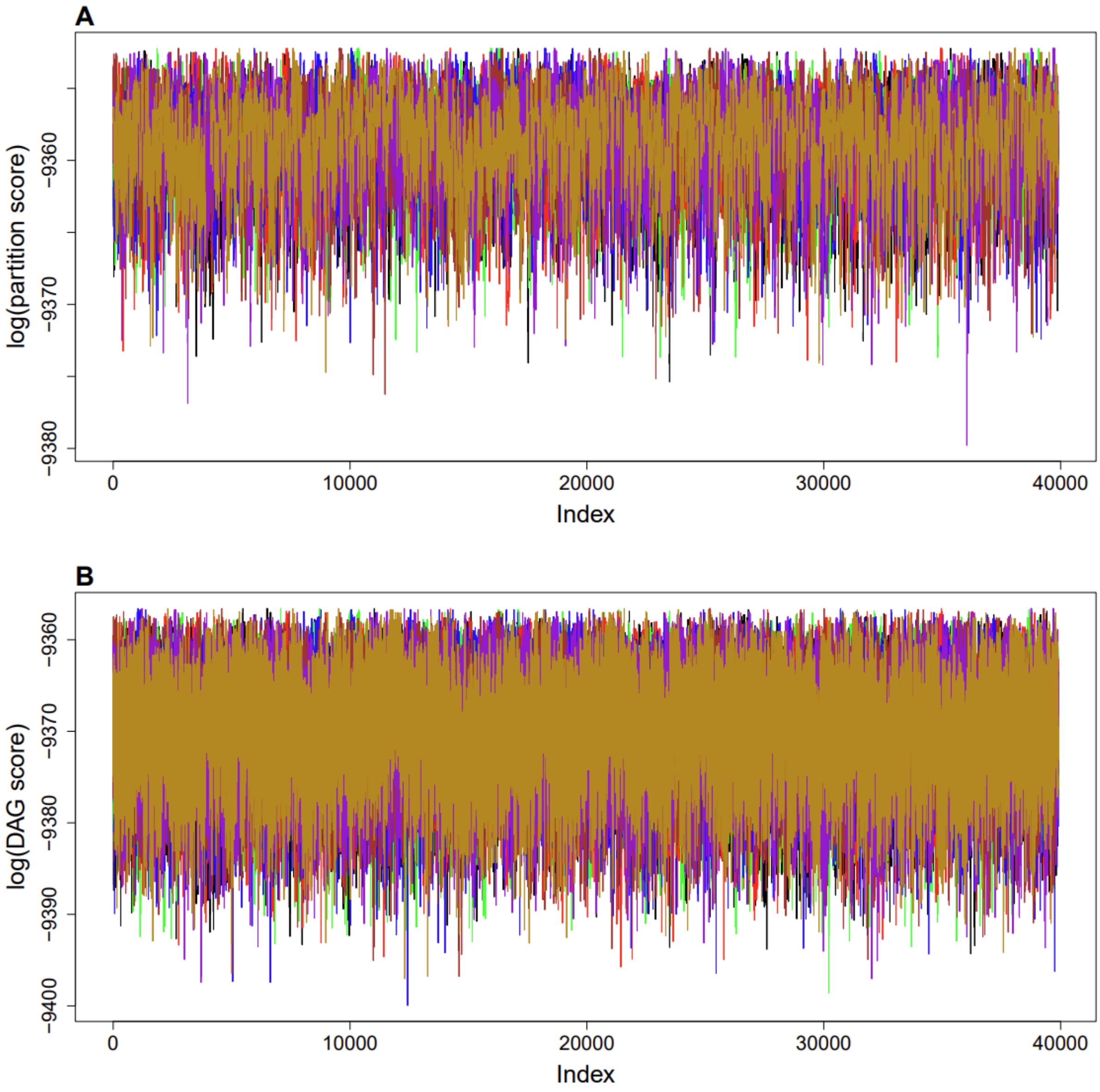

**Supplementary Fig. 4.|** The post burn-in log posterior trace for the chains in partition (A) and DAG (B) spaces, respectively. Each chain is represented by a different colour.

We also checked for the convergence of the marginalised edge probabilities using a concordance plot[3]. The marginalised edge probability for each pairwise edge (i.e., $p(A \rightarrow B|D)$) within each chain is calculated as the propensity of the edge appearing within that chain. The qualitative comparison across chains is shown in Supplementary Figure 5. We show that the marginalised edge probabilities across any two chains $c$ and $c'$ lie close to the $p_c(A \rightarrow B|D) = p_{c'}(A \rightarrow B|D)$ line. The range of the average absolute differences between any two chains given by,

$$\mu_{c,c'} = (1/n) \sum_{\{(A \rightarrow B)|(A \rightarrow B) \notin Q\}} |p_c(A \rightarrow B|D) - p_{c'}(A \rightarrow B|D))|$$

for all $n$ edges $(A \rightarrow B)$ that are not in the blacklist $Q$ were between 0.003 to 0.009.

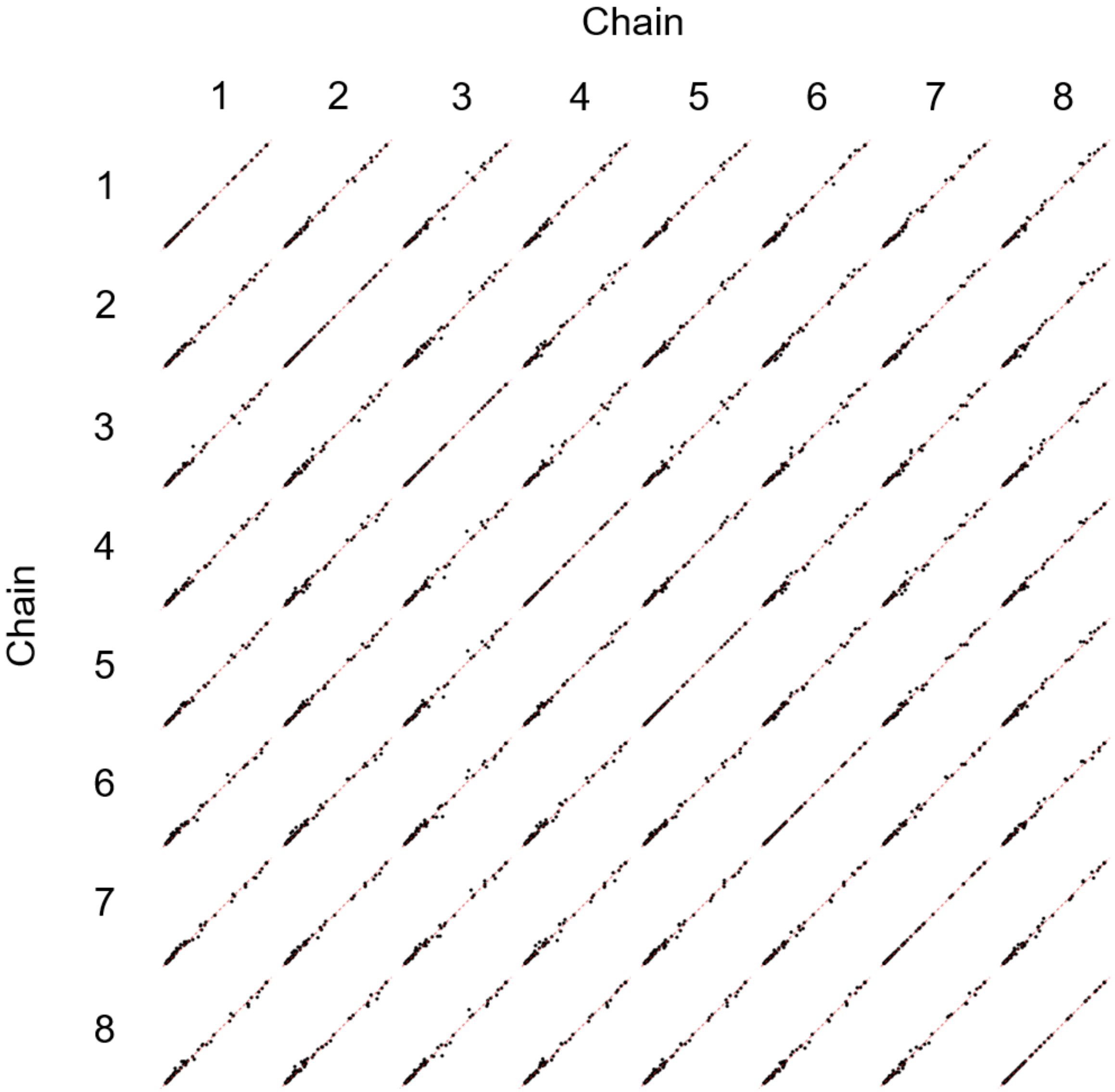

**Supplementary Figure 5.|** The comparison of marginalised edge probabilities across chains. Each point corresponds to the edge probability for chain $c$ compared to chain $c'$. The red dashed line corresponds to the edge probabilities being equal across two chains.

### Supplementary Note 6. Bayesian decision-theoretic framework

We take a Bayesian decision-theoretic approach to encode the decision-making process within our recommendation algorithm[4]. We define the $j$-th action $a_j \in \mathcal{A}$ to be a function that transforms a state $\boldsymbol{\theta}_i \in \boldsymbol{\Theta}$ for individual $i$ to their outcomes that will be observed at follow-up given by $\boldsymbol{Z}_i \in \mathcal{Z} = ('\text{poor}', '\text{fair}', '\text{healthy}')^n$ over the $n = 7$ domains. The components of $\boldsymbol{\theta}_i$ are the graph $G$, the parameters associated with that graph $\beta_G \in \mathcal{B} = [0, 1]^n$, and an individual's baseline observations $\boldsymbol{X}_i$, thus

$$\boldsymbol{Z}_i = a_j(\boldsymbol{\theta}_i) = a_j(G, \boldsymbol{\beta}_G, \boldsymbol{X}_i).$$

The utility function used to assign numeric values representing outcome preferences for individual $i$ given their outcomes $\boldsymbol{Z}_i$ is given by $U(\boldsymbol{Z}_i = a_j(G, \boldsymbol{\beta}_G, \boldsymbol{X}_i)) \in \mathbb{R}$. We define the multi-attribute utility function as a sum of sub-utility values across the domains, given by

$$U(\boldsymbol{Z}_i) = 7 \times \sum_{j=1}^{7} \frac{w_j u(Z_i^j)}{W}$$

where $u(Z_i^j) \in [0, 1]$ is the individual $i$'s sub-utility value for domain $j$. Given weakly-ranked preferences for domains defined in $\boldsymbol{h} = (h_1, h_2, \ldots, h_7)$ the unnormalized weight for the $j$-th domain is given by $w_j = (\sqrt{2})^{h_j}$ which is normalised by dividing by $W = \sum_{j=1}^{7} (\sqrt{2})^{h_j}$ [5,6]. The multiplication of the utility function by 7 maps the range back to [0, 7] with a change of a domain from 'poor' to 'healthy' with all else being equal corresponding to a change of 1 in the utility.

The actions $a_j(\boldsymbol{X}_i, G, \boldsymbol{\alpha}_G)$ span the action space $\mathcal{A}$ and are indexed by $j \in \{\emptyset, 1, \ldots, 7\}$ where $\emptyset$ corresponds to not selecting a domain (i.e., 'doing nothing') and the remaining indexes correspond to selecting the different domains as intervention targets. Outcomes under $j$ for $j \neq \emptyset$ are simulated from the Bayesian network defined by $(G, \boldsymbol{\beta}_G)$ after performing a do operation[7] of the form $do(Z_i^j = '\text{healthy}')$ and setting the baseline nodes consistent with the individual's observed values $\boldsymbol{X}_i$. Doing nothing corresponds to the action indexed by $\emptyset$, where we simulate outcomes using the BN after setting the baseline values to $\boldsymbol{X}_i$.

The optimal action is that which maximises the expected utility. Therefore, the utility must be averaged over the posterior distribution for $(\boldsymbol{Z}_i, G, \boldsymbol{\beta}_G)$ given all observations across the population $\boldsymbol{D} = \{\boldsymbol{X}, \boldsymbol{Z}\}$ given by $p_j(\boldsymbol{Z}_i, G, \boldsymbol{\beta}_G | \boldsymbol{D})$. The index $j$ on the posterior distribution is to recognise that the distribution for the outcomes changes under an action. The expected utility is then,

$$E_{p_j(\boldsymbol{Z}_i, G, \boldsymbol{\beta}_G | \boldsymbol{D})}[U(\boldsymbol{Z}_i)] = \sum_{G \in \mathcal{G}} \int_{\mathfrak{B}} \int_{\mathcal{Z}} U(\boldsymbol{Z}_i = a_j(\boldsymbol{X}_i, G, \boldsymbol{\beta}_G)) p_j(\boldsymbol{Z}_i, G, \boldsymbol{\beta}_G | \boldsymbol{D}) d\boldsymbol{Z}_i d\boldsymbol{\beta}_G.$$

Which can be put into the following form,

$$\begin{aligned}
E_{p_j(\boldsymbol{Z}_i, G, \boldsymbol{\beta}_G | \boldsymbol{D})}[U(\boldsymbol{Z}_i)] &= \sum_{G \in \mathcal{G}} \int_{\mathfrak{B}} \int_{\mathcal{Z}} U(\boldsymbol{Z}_i) p_j(\boldsymbol{Z}_i, G, \boldsymbol{\beta}_G | \boldsymbol{D}) d\boldsymbol{Z}_i d\boldsymbol{\beta}_G \\
&= \sum_{G \in \mathcal{G}} \int_{\mathfrak{B}} \int_{\mathcal{Z}} U(\boldsymbol{Z}_i) p_j(\boldsymbol{Z}_i | G, \boldsymbol{\beta}_G, \boldsymbol{D}) p(\boldsymbol{\beta}_G | G, \boldsymbol{D}) p(G | \boldsymbol{D}) d\boldsymbol{Z}_i d\boldsymbol{\beta}_G \\
&= \sum_{G \in \mathcal{G}} \int_{\mathfrak{B}} \left[ \int_{\mathcal{Z}} U(\boldsymbol{Z}_i) p_j(\boldsymbol{Z}_i | G, \boldsymbol{\beta}_G, \boldsymbol{D}) d\boldsymbol{Z}_i \right] p(\boldsymbol{\beta}_G | G, \boldsymbol{D}) p(G | \boldsymbol{D}) d\boldsymbol{\beta}_G \\
&= \frac{7}{W} \sum_{G \in \mathcal{G}} \int_{\mathfrak{B}} \left[ \sum_{j=1}^{7} E_{p_j(\boldsymbol{Z}_i | G, \boldsymbol{\beta}_G, \boldsymbol{D})} [w_j u(Z_i^j)] \right] p(\boldsymbol{\beta}_G | G, \boldsymbol{D}) p(G | \boldsymbol{D}) d\boldsymbol{\beta}_G
\end{aligned}$$

Using the gRain library we use a *maximum a posteriori* estimate of $\boldsymbol{\beta}_G$ for each graph denoted by $\widehat{\boldsymbol{\beta}}_G$, thus we are approximating the expectation of the utility as,

$$\begin{aligned}
E_{p_j(\boldsymbol{Z}_i, G, \boldsymbol{\beta}_G | \boldsymbol{D})}[U(\boldsymbol{Z}_i)] &\approx E_{p_j(\boldsymbol{Z}_i, G | \boldsymbol{D})}[U(\boldsymbol{Z}_i = a_j(\boldsymbol{X}_i, G, \widehat{\boldsymbol{\beta}}_G))] \\
&\approx \frac{7}{W} \sum_{G \in \mathcal{G}} \left[ \sum_{j=1}^{7} E_{p_j(\boldsymbol{Z}_i | G, \boldsymbol{D})} \left[ w_j u \left( Z_i^j = a_j(\boldsymbol{X}_i, G, \widehat{\boldsymbol{\beta}}_G)_i \right) \right] \right] p(G | \boldsymbol{D}).
\end{aligned}$$

Using the above approximation to the expectation of utility, the index for the optimal intervention target for individual $i$ is given by

$$j_i^* = \arg_j \max E_{p_j(\boldsymbol{Z}_i, G | \boldsymbol{D})}[U(\boldsymbol{Z}_i)].$$

We also report the standard error of the utility as,

$$SE_{p_j(\boldsymbol{Z}_i,G,\boldsymbol{\beta}_G|\boldsymbol{X})}[U(\boldsymbol{Z}_i)] \approx \sqrt{Var(E_{p_j(\boldsymbol{Z}_i,G|\boldsymbol{X})}[U(\boldsymbol{Z}_i)])}$$

which only considers the contribution to the variance from the uncertainty over the graph. The average treatment effects effect for individual $i$ given an intervention on $j$ is estimated by,

$$ATE_i^j \approx E_{p_j(\boldsymbol{Z}_i,G|\boldsymbol{X})}[U(\boldsymbol{Z}_i)] - E_{p_\emptyset(\boldsymbol{Z}_i,G|\boldsymbol{X})}[U(\boldsymbol{Z}_i)].$$

### Supplementary Note 7. Linear causal effects

In addition to the main analysis, where we assumed non-parametric relationships between variables, we also estimated the linear causal effects, to understand the typical change in one domain given an intervention on another. The linear causal effect of $X$ on $Y$ given a DAG $G$ is denoted by $\theta_{X,Y,G}$, and is the linear gradient when $X$ is an ancestor of $Y$ adjusted for a control set and 0 otherwise. A sufficient control set that blocks the backdoor paths from $X$ to $Y$ is the parents of $X$ given a DAG $G$ denoted by $\boldsymbol{Z}_{X,G}$ [8]. As such, the linear causal effect for a given graph $G$ is then,

$$\theta_{X,Y,G} = \begin{cases} \beta_{X,Y}|\boldsymbol{Z}_{X,G} \text{ if } X \text{ is an ancestor of } Y \text{ in } G \\ 0 \text{ otherwise} \end{cases}$$

where we assume the linear model,

$$Y = \beta_{X,Y}X + \boldsymbol{\gamma}^{\top}\boldsymbol{Z}_{X,G}.$$

with $\boldsymbol{\gamma}$ being the linear regression coefficients and we simplify our notation such that $\beta_{X,Y} = \beta_{X,Y}|\boldsymbol{Z}_{X,G}$.

We then endeavour to estimate the posterior distribution for the causal effect marginalised over the graph structure. The posterior distribution for the marginalised causal effect $\Phi_{X,Y}$ is given by,

$$p(\Phi_{X,Y}|D) = \Sigma_G p(\theta_{X,Y,G}|G)p(G|D).$$

Similar to the main analysis, we make an approximation to the marginalised causal effect by using an *maximum a posteriori* estimate for $\beta_{X,Y}$ that we denote as $\hat{\beta}_{X,Y}$, with the causal effect denoted as $\hat{\theta}_{X,Y,G}$. The approximate posterior distribution is then,

$$p(\Phi_{X,Y}|D) \approx \Sigma_G p(\hat{\theta}_{X,Y,G}|G)p(G|D).$$

Similar to the main analysis, the bulk of the uncertainty is in the graph structure, and therefore we consider this to be a reasonable approximation to the posterior distribution. We provide a summary of the posterior distribution for the linear causal effects in Supplementary Table 8. Intuitively, the linear causal effect provides an estimate for the change in $X$ given a one-point change in $Y$ where 'poor', 'fair', and 'healthy' are equally spaced by one-point.

| | Functioning | Distress | Nutrition | Physical Activity | Sleep | Social | Substance |
|---|---|---|---|---|---|---|---|
| Functioning | 1 | 0.02 (0, 0.19) | 0 (0, 0.09) | 0 (-0.02, 0.07) | 0.01 (0, 0.12) | 0.01 (0, 0.14) | 0 (0, 0.05) |
| Distress | 0.27 (0, 0.46) | 1 | 0.03 (-0.02, 0.17) | 0.12 (0, 0.23) | 0.28 (0, 0.40) | 0.28 (0, 0.41) | 0 (-0.05, 0.03) |
| Nutrition | 0 (0, 0.07) | 0 | 1 | 0 (0, 0.10) | 0 (0, 0.03) | 0 (0, 0.11) | 0 (-0.01, 0) |
| Physical Activity | 0 (0, 0.04) | 0 (0, 0.09) | 0 (0, 0.08) | 1 | 0 (0, 0.13) | 0 (0, 0.12) | 0 (-0.04, 0) |
| Sleep | 0.01 (0, 0.18) | 0.03 (0, 0.41) | 0 (-0.07, 0.05) | 0.13 (0, 0.25) | 1 | 0.04 (0, 0.28) | 0 (-0.03, 0.01) |
| Social | 0.01 (0, 0.13) | 0.01 (0, 0.20) | 0.01 (0, 0.11) | 0.03 (0, 0.15) | 0.04 (0, 0.17) | 1 | 0 (-0.05, 0) |
| Substance | 0 (0, 0.09) | 0 | 0 (-0.04, 0.00) | 0 (-0.09, 0) | 0 (0, 0.02) | 0 (-0.06, 0) | 1 |

**Supplementary Table 8|** The mean (95% equal-tailed credible intervals) for the linear causal effects from follow-up to follow-up. Where the pathway is defined as row → column.
